# The intersection between migration, HIV, and family planning in Uganda: a cross-sectional population-based study

**DOI:** 10.1101/2023.08.05.23293691

**Authors:** Prossy Namusisi, Ping Teresa Yeh, Robert Ssekubugu, Larry William Chang, Tom Lutalo, Linnea A. Zimmerman, Mary Kathryn Grabowski

## Abstract

**Background:** Low use of modern methods of contraception has been linked to HIV seropositivity and to migration, but few studies have evaluated the intersection of both risk factors with contraceptive use.

**Methods:** We analyzed cross-sectional data from sexually active female participants aged 15 to 49 years in the Rakai Community Cohort Study (RCCS) between 2011 and 2013. The RCCS is an open population-based census and individual survey in south-central Uganda. Recent in-migrants (arrival within approximately 1.5 years) into RCCS communities were identified at time of household census. The primary outcome was unsatisfied demand for a modern contraceptive method (injectable, oral pill, implant, or condom), which was defined as non-use of a modern contraceptive method among female participants who did not want to become pregnant in the next 12 months. Poisson regression models with robust variance estimators were used to identify associations and interactions between recent migration and HIV serostatus on unsatisfied contraceptive demand.

**Results:** There were 3,417 sexually active participants with no intention of becoming pregnant in the next year. The mean age was 30 (±8) years, and 17.3% (n=591) were living with HIV. Overall, 43.9% (n=1,500) were not using any modern contraceptive method. Recent in-migrants were somewhat more likely to have unsatisfied contraceptive demand as compared to long-term residents (adjusted prevalence risk ratio [adjPRR]=1.14; 95% confidence interval [95%CI]: 1.02–1.27), whereas participants living with HIV were less likely to have unsatisfied contraceptive demand relative to HIV-seronegative participants (adjPRR=0.80; 95%CI=0.70-0.90). When stratifying on migration and HIV serostatus, we observed the highest levels of unsatisfied contraceptive demand among in-migrants living with HIV (48.7%); however, in regression analyses, interaction terms between migration and HIV serostatus were not statistically significant.

**Conclusions:** Unsatisfied contraceptive demand was high in this rural Ugandan setting. Being an in-migrant, particularly among those living with HIV, was associated with higher unsatisfied contraceptive demand.

## Plain English summary

Through a cross-sectional study, we explored the relationship between HIV status, migration, and contraceptive use among sexually active women of reproductive age in rural south-central Uganda. People who had moved into the study area within the last 1.5 years were considered in-migrants, compared to long-term residents i.e. people who had not moved. We examined unsatisfied demand for a modern contraceptive method, which is to say female participants who did not want to become pregnant in the next 12 months and were not using at least one of the following contraceptive methods: injectable, oral pill, implant, or condom. We included 3,417 sexually active female participants with no intention of becoming pregnant in the next year. The average age of these women was 30 years, less than 20% were living with HIV, and almost half were not using any modern contraceptive methods. Recent in-migrants were somewhat more likely to have unsatisfied contraceptive demand as compared to long-term residents, whereas participants living with HIV were less likely to have unsatisfied contraceptive demand relative to HIV-negative participants. Being an in-migrant, particularly among those living with HIV, was associated with higher unsatisfied contraceptive demand. This study shows the need for integrating family planning and HIV services for mobile populations in East Africa.

## Background

Addressing women’s demand for contraceptive services can prevent unintended pregnancies (**1, 2**) and reduce mother-to-child HIV transmission among women living with HIV (**3**). In recognition of the importance of contraception as a sexual and reproductive health intervention, total demand satisfied by a modern contraceptive method (the percentage of women 15-49 years who are currently using a modern method of contraception among women 15-49 who wish to prevent or delay pregnancy), was included in the Sustainable Development Goals, with the goal of satisfying 75% of demand by 2035 (4). Despite this ambitious goal, much work remains. Family Planning 2030 recently estimated that 19.7% of all women in sub-Saharan Africa as of 2022 have unmet need for contraception (5) – that is, they state a desire to delay or prevent pregnancy but are not using a method of contraception. Recent research highlights persistent equity gaps in meeting demand for family planning services across sub-Saharan Africa (6). While equity gaps have been documented across multiple sociodemographic characteristics, including age, wealth, and education status (6–9), there are relatively fewer studies evaluating how demand satisfied varies among women living with HIV.

Demand satisfied with a modern method is increasingly recognized as a better measure to track progress towards meeting family planning goals than prevalence estimates of unmet need or contraceptive use, as estimates of both unmet need and the contraceptive prevalence rate include women who are not at risk for unintended pregnancy (e.g., women who are pregnant or who are not sexually active) in the denominator. In contrast, demand satisfied is specific to women who wish to delay or prevent childbearing and, thus, provides a better measure of the ability of family planning programs to meet the needs of women who may benefit from the use of contraception based on their sexual activity and fertility intentions. These considerations are particularly pertinent among women living with HIV, as HIV status has been demonstrated to affect fertility intentions (10, 11), and in turn, demand for contraception. HIV-positive serostatus has been linked to greater unmet need for contraception in multiple settings across the African continent (12–15), although many of these prior studies employ facility-based samples that limit generalizability to the broader population and were conducted before widespread availability of antiretroviral therapy (ART) (15). Since the early 2000s, access to ART has gradually increased in sub-Saharan Africa, resulting in higher levels of HIV viral load suppression among women living with HIV (16, 17). ART scale-up has also occurred during a period of wide-spread commitment to improve access to contraceptive services (18), including among women living with HIV (19). Little research, however, has explored whether improvements in access to services and increases in contraceptive use in the general population have resulted in improved equity in demand satisfied by HIV serostatus.

Additionally, prior research has additionally shown that migration is a risk factor for HIV acquisition and that women living with HIV are more likely to migrate. Migration can also affect contraceptive need by changing (either increasing or decreasing) sexual exposure between partners through absence from home (20, 21), and through exposing migrants to differing fertility norms (22), generally leading towards lower desired fertility and greater need for contraception. The relationship between contraceptive behavior for contraception and migration is complex and depends in part on both a migrant’s place of origin and destination. For example, one cross-sectional study conducted in Zambia found significantly greater likelihood of experiencing unmet need for contraception among rural non-migrants compared to urban non-migrants and rural-to-rural migrants, but not other groups (i.e. urban-to-rural migrants, urban-to-urban migrants, rural-to-urban migrants) (23). Another cross-sectional study in Ethiopia found that unmet need for contraception was significantly lower among rural-to-urban migrants compared to rural non-migrants (24). In both studies, however, the reference group included women who desired to have children, were pregnant, and/or were not sexually active. It is thus not possible to differentiate between the effect of inequitable access to and use of services or differences in fertility intentions and sexual behavior. As with HIV, however, few studies have explored whether satisfied contraceptive demand, which addresses this limitation, differs between migrants and non-migrants despite the influence that migration may play on both the demand for contraception and ability of migrants to access health services. In addition to changes in contraceptive access, migration may also lead to interruption in continuity of care among women living with HIV (25), which may include their family planning services. HIV-associated stigma may also hinder migrating women living with HIV from accessing both HIV and family planning services at their place of destination (26). Despite strong associations between migration and HIV, and the impact of both on use of family planning services, there is very limited research on how these factors interact to influence total demand satisfied by a contraceptive method among African women.

Here, we used data from the Rakai Community Cohort Study (RCCS) in south-central Uganda to assess contraceptive use patterns among sexually active women with no intention of getting pregnant in the next 12 months living with and without HIV. Our objective was to determine whether the relationship between a recent history of migration and demand satisfied varies by HIV serostatus. Given the large population of women living with HIV, increasing migration as countries urbanize, and the ongoing development of dual HIV prevention and family planning technologies, understanding the relationship between unsatisfied demand for contraception, migration, and HIV in sub-Saharan Africa is critical to effective delivery of integrated HIV and family planning services.

## Methods

### Study setting and population

We used data from 30 rural agrarian and semi-urban trading communities under surveillance during the fifteenth survey round (R15) of the RCCS, collected between July 2011 and May 2013. The RCCS is a longitudinal open population-based household census and behavioral survey of persons aged 15 to 49 years in south-central Uganda conducted at approximately 18-month intervals. During the RCCS household census, data on in- and out-migration, births, and deaths is obtained (27). Almost all the migrations are internal, as opposed to cross-border or international, migration. At the time of RCCS survey, data on participant demographics, sexual behaviors and partnerships, and health service utilization, including use of family planning methods, are obtained. Though the RCCS is currently ongoing, RCCS survey R15 was the most recent survey visit for which comprehensive data on fertility intention and contraceptive method use were obtained.

### Exposure variables

Our primary exposure variables were migration and HIV serostatus. Migration status was a binary exposure variable with participants being classified as either a recent in-migrant or a long-term resident. A recent in-migrant was defined as a woman who moved into an RCCS study community after the most recent previous RCCS household census (RCCS R14, approximately 1.5 year interval between RCCS R14 and R15) with intention to stay in the community. In-migrants were identified at time of household census, which took place approximately 4 weeks before the RCCS survey. Conversely, long-term residents were defined as women with no recent history of migration over the preceding survey interval (27). HIV serostatus was measured using a validated three-rapid-test (Determine, Stat Pak, and Unigold) algorithm on serum samples collected at time of RCCS survey and confirmed with enzyme-linked immunosorbent assays at the Rakai Health Sciences Program laboratories in Kalisizo, Uganda as previously described (28).

### Primary outcome

Women were surveyed as to whether they were using any modern methods of contraception during the last 12 months. Specifically, they were queried, one by one, on whether they were currently using the following modern contraceptive methods: combined oral contraceptive pill, condom, Depo-Provera, intra-uterine device (IUD)/coil, implant/Norplant, or bilateral tubal ligation. They were also asked if they had any intention of becoming pregnant in the next 12 months. Our primary outcome was unsatisfied demand for a modern contraceptive, which we defined as non-use of modern contraceptive method at the time of the survey among sexually active women in the RCCS with no intention of becoming pregnant in the next 12 months.

### Statistical analysis

Women were classified into four groups: (1) HIV-seropositive and recent in-migrant, (2) HIV-seropositive and long-term resident, (3) HIV-seronegative and recent in-migrant, and (4) HIV-seronegative and long-term resident. For each of these four groups, we tabulated their frequency and percent of various sociodemographic characteristics: age (15-24, 25-34, 35-49 years), education level (some primary, post-primary), marital status (never married, currently married, previously married), socio-economic status (lowest, low-middle, high-middle, highest as determined using a locally validated scale)(29), religion (Muslim, Catholic, Protestant/Pentecostal, other), and occupation (agriculture/housework, non-agriculture i.e. bar/restaurant/salon, trade/shopkeeper, government/teacher) and current ART use (yes, no).

We then tabulated the use of different types of contraceptive methods (including non-use of any modern methods) by migration and HIV serostatus. We also conducted sensitivity analysis excluding condoms, since condoms are a commonly used method of HIV prevention and tend to have lower efficacy for pregnancy prevention in our study setting as compared to other modern contraceptive methods (30, 31). Further, we disaggregated unsatisfied contraceptive demand by age (<30 years versus ≥ 30 years), since it previously has been associated with younger age among individuals living with HIV (15).

Poisson regression with robust variance was used to estimate unadjusted and adjusted prevalence risk ratios (PRR) and 95% confidence intervals (95% CI) of unsatisfied contraceptive demand by migration history and HIV serostatus. Based on our conceptual framework and prior literature review, we included the following as potential confounders in our multivariable model: age, education, marital status, socio-economic status, religion, and occupation. To assess for potential effect modification, we stratified analyses by HIV and migration status included product (i.e., interaction) terms between HIV and migration status in regression models, which were considered statistically significant at an alpha significance level of 0.05. All analyses were conducted using STATA (version BE17; College Station, Texas).

### Ethical review

This study was approved by the Western Institutional Review Board and the Research Ethics Committee of the Uganda Virus Research Institute. The study received clearance by the Ugandan National Council for Science and Technology. All study participants provided written informed consent or assent/parental consent for participants aged less than 18 years old.

## Results

There were 10,960 individuals residing in RCCS rural and semi-urban trading communities who participated in RCCS R15 including 6,083 women of whom 3,417 were sexually active women and did not want to become pregnant in the next year. Of these 3,417 women, 2,346 (68.7%) were long-term residents who were HIV-seronegative, 476 (13.93%) were long-term residents living with HIV, 480 (14.1%) were recent in-migrants who were HIV-seronegative, and 115 (3.37%) were recent in-migrants living with HIV. Table 1 presents sociodemographic characteristics of the study participants. The majority were 25–34 years of age, currently married, and with some primary education. Women living with HIV tended to be older, less likely to be married, and have lower SES compared to women who were HIV seronegative, while recent in-migrants tended to be younger and employed in occupations outside of agriculture and housework. Of those living with HIV, 54% reported current ART use, although levels of ART use were higher among long-term residents as compared to recent migrants living with HIV.

**Table 1.** Characteristics of RCCS female study participants by recent migration history and HIV serostatus.

| **Participant characteristics** | **Long-term resident HIV-seronegative N (%)** | **Long-term resident HIV-seropositive N (%)** | **Recent in-migrant HIV-seronegative N (%)** | **Recent in-migrant HIV-seropositive N (%)** | **Total N (%)** |
| --- | --- | --- | --- | --- | --- |
| **Total** | 2346 (68.7) | 476 (13.9) | 480 (14.0) | 115 (3.4) | 3417 (100%) |
| **Age (years)** |  |  |  |  |  |
| 15-24 | 561 (23.9) | 56 (11.8) | 255 (53.1) | 38 (33.0) | 910 (26.6) |
| 25-34 | 1022 (43.6) | 203 (42.7) | 183 (38.1) | 54 (47.0) | 1462 (42.8) |
| 35-49 | 763 (32.5) | 217 (45.6) | 42 (8.8) | 23 (20.0) | 1045 (30.6) |
| **Education** |  |  |  |  |  |
| Primary or none | 1382 (58.9) | 310 (65.1) | 228 (47.5) | 71 (61.7) | 1991 (58.3) |
| Post primary | 964 (41.1) | 166 (34.9) | 252 (52.5) | 44 (38.3) | 1426 (41.7) |
| **Marital status** |  |  |  |  |  |
| Never married | 339 (14.5) | 41 (8.6) | 72 (15.0) | 11 (9.6) | 463 (13.6) |
| Currently married | 1735 (74.0) | 282 (59.2) | 336 (70.0) | 58 (50.4) | 2411 (70.6) |
| Previously married | 272 (11.6) | 153 (32.1) | 72 (15.0) | 46 (40.0) | 543 (15.9) |
| **SES** |  |  |  |  |  |
| Lowest | 326 (13.9) | 68 (14.3) | 38 (7.9) | 12 (10.4) | 444 (13.0) |
| Low middle | 696 (29.7) | 147 (30.9) | 129 (26.9) | 44 (38.3) | 1016 (29.8) |
| High middle | 620 (26.5) | 156 (32.8) | 166 (34.6) | 37 (32.2) | 979 (28.7) |
| Highest | 702 (30.0) | 105 (22.1) | 147 (30.6) | 22 (19.1) | 976 (28.6) |
| **Religion** |  |  |  |  |  |
| Other | 18(0.8) | 0 (0.0) | 5 (1.0) | 1 (0.9) | 24 (0.7) |
| Catholic | 1578 (67.3) | 312 (65.6) | 271 (56.5) | 69 (60.0) | 2230 (65.3) |
| Protestant/Pentecostal | 438 (18.7) | 104 (21.9) | 129 (26.9) | 33 (28.7) | 704 (20.6) |
| Muslim | 312 (13.3) | 60 (16.6) | 75 (15.6) | 12 (10.4) | 459 (13.4) |
| **Occupation** |  |  |  |  |  |
| Agriculture/housework | 1444 (61.6) | 281 (59.0) | 223 (46.5) | 49 (42.6) | 1997 (58.4) |
| Non agriculture | 902 (38.5) | 195 (41.0) | 257 (53.5) | 66 (57.4) | 1420 (41.6) |
| **Self-reported ART use** |  |  |  |  |  |
| Currently on ART | 0 (0.0) | 194 (60.8) | 0 (0) | 19 (47.5) | 213 (53.7) |
| Not currently on ART | 30 (100) | 125 (39.2) | 8 (100) | 21 (52.5) | 184 (46.4) |

Table 2 shows the different methods of modern contraceptive use overall and stratified by migration history and HIV serostatus. Depo-Provera was the most commonly used modern contraceptive method (29.4%), followed by condoms (13.7%). Women living with HIV were more likely to report using condoms, irrespective of migration status. Overall, unsatisfied contraceptive demand was high, regardless of migration or HIV status. Recent in-migrants living with HIV had the highest levels of unsatisfied contraceptive demand (48.7%), while long-term residents living with HIV had the lowest levels of unsatisfied contraceptive demand (36.6%); however, lower levels of unsatisfied contraceptive demand observed among long-term residents living with HIV was entirely driven by condom use.

**Table 2.** Modern contraceptive use, overall and by recent migration history and HIV serostatus.

| **Modern contraceptive method** | **Long-term resident HIV-seronegative N (%)** | **Long-term resident HIV-seropositive N (%)** | **Recent in-migrant HIV-seronegative N (%)** | **Recent in-migrant HIV-seropositive N (%)** | **Total N (%)** |
| --- | --- | --- | --- | --- | --- |
| Pill | 138 (5.9) | 14 (2.9) | 21 (4.4) | 2 (1.7) | 175 (5.1) |
| Condom | 297 (12.7) | 99 (20.8) | 53 (11.0) | 19 (16.5) | 468 (13.7) |
| Depo-Provera | 690 (29.4) | 148 (31.1) | 138 (28.8) | 30 (26.1) | 1006 (29.4) |
| IUD/coil | 48 (2.1) | 9 (1.9) | 10 (2.1) | 2 (1.7) | 69 (2.0) |
| Implant/Norplant | 113 (4.8) | 28 (5.9) | 28 (5.8) | 5 (4.4) | 174 (5.1) |
| Bilateral tubal ligation | 31 (1.3) | 8 (1.6) | 0 (0.0) | 1 (0.9) | 40 (1.1) |
| Unsatisfied contraceptive demand* | 1038 (44.3) | 174 (36.6) | 232 (48.3) | 56 (48.7) | 1500 (43.9) |
| Unsatisfied contraceptive demand, excluding condoms* | 1326 (56.5) | 269 (56.6) | 283 (59.0) | 75 (65.2) | 1953 (57.2) |

\*Unsatisfied contraceptive demand was defined non-use of a modern contraceptive method; in sensitivity analysis, condoms were excluded from modern contraceptive methods.

Figure 1 shows unsatisfied contraceptive demand by HIV, migration status, and by age. For women both below and above 30 years, in-migrants had higher levels of unsatisfied contraceptive demand as compared to long-term residents. However, older women tended to have higher unsatisfied contraceptive demand. Overall, in-migrants living with HIV had the highest levels of unsatisfied contraceptive demand.

**Figure 1.**
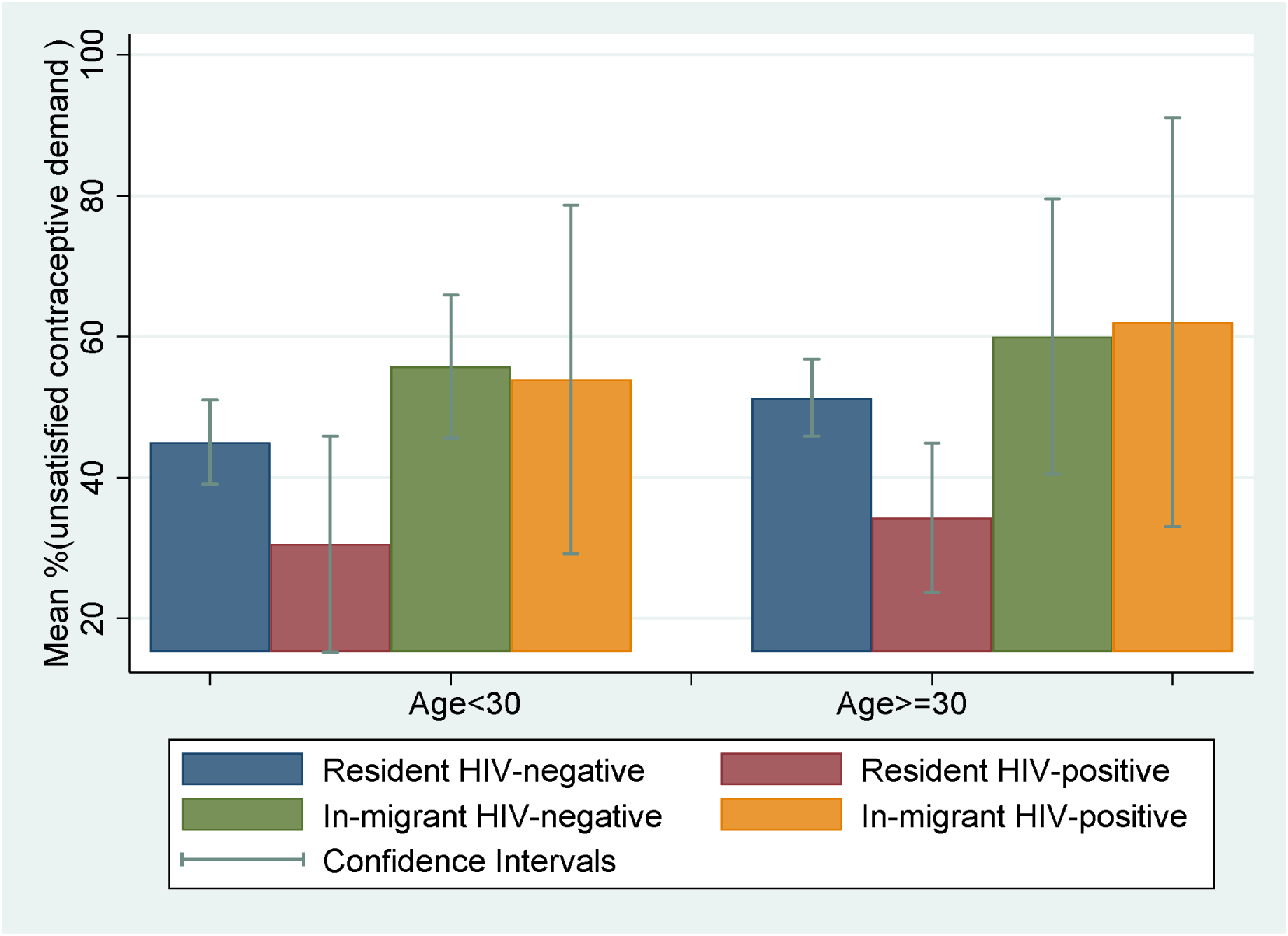
Prevalence of unsatisfied contraceptive demand by recent migration history and HIV serostatus stratified by age among 3,417 sexually active females with no intention of getting pregnant in the next year.

Table 3 shows unadjusted and adjusted associations between HIV and migration and unsatisfied contraceptive demand. Participants living with HIV were significantly less likely to have unsatisfied demand as compared to HIV-seronegative women (adjPRR=0.80; 95% CI 0.71–0.91); however, the lower levels of unsatisfied demand among those living with HIV were entirely driven by higher levels of condom use in this group (Supplemental Table 1). Conversely, recent in-migrants had a 14% higher prevalence of unsatisfied contraceptive demand as compared to long-term residents (adjPRR=1.14; 95% CI 1.02–1.27). There was some evidence of a positive interaction between HIV serostatus and recent migration, such that the impact of migration on unsatisfied contraceptive demand was greater among participants living with HIV; however, results were not statistically significant (interaction p-value=0.096). Furthermore, among participants living with HIV, unsatisfied contraceptive demand was 61.0% among participants who self-reported ART use and 39.0% among those who did not (PRR=1.07 95% CI 0.80-1.44). Older female age, lower SES, and agricultural work were all associated with higher levels of unsatisfied contraceptive demand.

**Table 3.** Factors associated with unsatisfied contraceptive demand among 3,417 sexually active female participants in the RCCS with no intention of getting pregnant in the next year.

|  | n/N (%) | Unadjusted PR<br>[95% CI] | p-value | Adjusted PR<br>[95% CI] | p-value |
| --- | --- | --- | --- | --- | --- |
| **Migration status** |  |  |  |  |  |
| Resident | 1212/2822 (43.0%) | Ref |  | Ref |  |
| In-migrant | 288/595 (48.4%) | 1.13 [1.03, 1.24] | 0.012 | 1.14 [1.02, 1.27] | 0.019 |
| **HIV serostatus** |  |  |  |  |  |
| HIV-seronegative | 1270/2826 (44.9%) | Ref |  | Ref |  |
| HIV-seropositive | 230/591 (38.9%) | 0.87 [0.78, 0.97] | 0.01 | 0.80 [0.70, 0.90] | <0.001 |
| **Interaction term between migration and HIV-positive serostatus** |  | 1.11 [0.92, 1.35] | 0.271 | 1.23 [0.96, 1.57] | 0.096 |
| **Age** |  |  |  |  |  |
| 15-24 | 416/910 (45.7%) | Ref |  | Ref |  |
| 25-34 | 578/1462 (39.5%) | 0.86 [0.79, 0.95] | 0.003 | 0.91 [0.82, 1.00] | 0.069 |
| 35-49 | 506/1045 (48.4%) | 1.06 [0.96, 1.64] | 0.233 | 1.12 [1.00, 1.25] | 0.036 |
| **Education** |  |  |  |  |  |
| Some primary | 894/1991 (44.9%) | Ref |  | Ref |  |
| Post primary | 606/1426 (42.5%) | 0.95 [0.88, 1.02] | 0.164 | 0.99 [0.91, 1.07] | 0.800 |
| **Marital status** |  |  |  |  |  |
| Never married | 208/463 (44.9%) | Ref |  | Ref |  |
| Currently married | 1040/2411 (43.1%) | 0.96 [0.86, 1.07] | 0.472 | 0.93 [0.82, 1.05] | 0.236 |
| Previously married | 252/543 (46.4%) | 1.03 [0.90, 1.18] | 0.638 | 0.99 [0.85, 1.16] | 0.945 |
| **SES** |  |  |  |  |  |
| Lowest | 224/444 (50.5%) | Ref |  | Ref |  |
| Low middle | 458/1016 (45.1%) | 0.89 [0.80, 1.00] | 0.054 | 0.91 [0.81, 1.02] | 0.096 |
| High middle | 438/979 (44.7%) | 0.89 [0.79, 1.00] | 0.042 | 0.92 [0.81, 1.03] | 0.144 |
| Highest | 379/976 (38.8%) | 0.77 [0.68, 0.87] | <0.001 | 0.80 [0.71, 0.91] | 0.001 |
| **Religion** |  |  |  |  |  |
| Other | 10/24 (41.7%) | Ref |  | Ref |  |
| Catholic | 954/2230 (42.8%) | 1.03 [0.64, 1.65] | 0.913 | 1.11 [0.68, 1.86] | 0.705 |
| Protestant/Pentecost | 325/704 (46.2%) | 1.11 [0.69, 1.79] | 0.676 | 1.20 [0.71, 2.03] | 0.495 |
| Muslim | 211/459 (46.0%) | 1.10 [0.68, 1.79] | 0.690 | 1.19 [0.70, 2.02] | 0.510 |
| **Occupation** |  |  |  |  |  |
| Agriculture/housework | 928/1997 (46.5%) | Ref |  | Ref |  |
| Non agriculture | 572/1420 (40.3%) | 0.87 [0.80, 0.94] | <0.001 | 0.87 [0.80, 0.95] | 0.002 |

PRR=Prevalence risk ratio; adjRR=adjusted prevalence risk ratio; 95% CI=95% confidence interval; Ref=reference

## Discussion

In this cross-sectional study assessing how the prevalence of non-use of contraception differed among recent in-migrants and long-term residents by HIV status in rural south-central Uganda, we observed that unsatisfied contraceptive demand among sexually active female participants with no intention of getting pregnant in the next year was 43.9% (including condoms, or 57.2% excluding condoms). We found that unsatisfied demand was significantly higher among recent in-migrants compared to long-term residents, and that women living with HIV had a 20% lower risk of unsatisfied contraceptive demand as compared to HIV-seronegative women, largely due to increased use of condoms among those with HIV. Our interaction analyses further suggested possible effect modification by migration on HIV, such that the impact of migration on unsatisfied demand was greater among those with HIV.

We have previously reported that migrating girls and women in Uganda tend to have higher levels of HIV acquisition risk and lower levels of ART use if living with HIV (32). We similarly found lower levels of ART use among female participants living with HIV in this study. In addition, we report that recent in-migrants also tend to have significantly higher levels of unsatisfied contraceptive demand. Taken together, these data suggest that migrating persons may generally have reduced access to health services in Uganda and that family planning and HIV programs in Uganda should consider the unique needs of mobile persons. Emerging long-acting dual prevention HIV and family technologies may be one way to help close gaps in unsatisfied contraceptive demand and HIV risk during periods of migration (33). However, unlike migration and ART use, prior literature on the association between migration and unsatisfied contraceptive demand from Africa, although very limited, is mixed, suggesting the impact of migration likely depends heavily on context (23, 24). Of note, most prior studies have examined the impact of migration by where a person is migrating to or from (i.e., rural to rural vs. rural to urban), which we did not do here. In addition, previous studies have included women with no need for contraception in their denominators, so cannot determine whether the observed unmet need is due to differences in fertility intentions or other factors. Thus, additional research is needed to disentangle the exact mechanisms by which migration may affect access to family planning services across a range of different settings and migration contexts, including reasons for migration (e.g., marriage).

Condom use was associated with lower levels of unsatisfied contraceptive demand among women living with HIV in this study. In addition to preventing pregnancy, condoms also prevent HIV transmission. However, condoms have lower levels of typical use effectiveness for pregnancy prevention as compared to other modern contraceptive methods (34), which increases the risk of unintended pregnancy when used as the only method of contraception. When excluding condoms from modern contraceptive methods, we observed no association between HIV serostatus and unsatisfied contraceptive demand. Higher levels of condom use among participants living with HIV may be because these women are more motivated to use condoms for both family planning and prevention of HIV transmission to their partners and because of better access to family planning through HIV services. A recent study of modern contraceptive use in Ghana found that women who recently tested for HIV were more likely to report using modern contraceptives (35). While limited access to condoms through HIV services during migration may partly explain the high levels of unsatisfied contraceptive demand among recently migrating women living with HIV, we also observed lower levels of all other modern contraceptive methods among this subgroup relative to long-term residents also living with HIV.

We also found that having higher SES was associated with lower prevalence of unsatisfied contraceptive demand. Disparities in contraceptive use and demand satisfied have been identified across a range of sociodemographic characteristics, including wealth and education (6, 7, 36). Disparities by wealth are particularly problematic, as wealth is generally highly correlated with other socioeconomic characteristics, such as residence and education (6), and disparities by wealth thus frequently indicate the presence of other inequities. This is reflected by our observation that women working in agriculture, who tend to be of lower SES and live in rural settings, also had significantly higher levels of unsatisfied contraceptive demand. Disparities by socioeconomic status likely exist for a number of reasons, from lower accessibility of services due to limited economic resources (37), lower quality counseling (38), and restricted autonomy (39) to make independent decisions related to contraceptive use relative to wealthier women (40). Lastly, consistent with prior studies, we observed higher levels of unsatisfied contraceptive demand among older women, which likely reflects lower perceived risk of pregnancy (41, 42), though research on the unique contraceptive needs of women over age 35 in Africa is limited.

This study has some important limitations. First, this was cross-sectional study. Consequently, were unable to assess temporal or causal relationships between migration and HIV on unsatisfied contraceptive demand. Second, this study was done in a small number of communities in southcentral Uganda, so results may not be generalizable to other populations. Relatedly, migration events captured in this study were to a predominantly rural area (RCCS communities), and so results may not be applicable to populations migrating to urban environments or migration across country borders. Fourth, the data for this study were collected more than a decade ago, and may not reelect modern relationships between HIV, migration, and modern contraceptive use. Finally, our sample size of participants living with HIV and recently migrating was limited and may have affected our ability to detect significant interactions between HIV and migration. Despite these limitations, this study has several notable strengths, including its population-based study design and restriction to women who were not intending to get pregnant in the next year. To the best of our knowledge, this is the first study in sub-Saharan Africa to evaluate the interaction between HIV and migration on unsatisfied contraceptive demand. Integrating family planning services and HIV services – and exploring implementation strategies for such integration – may be of great benefit to women living with HIV in this context, especially for mobile populations.

## Conclusion

Unsatisfied modern contraceptive demand is high among girls and women in rural southcentral Uganda, particularly among persons with a recent history of migration. Impact of migration on use of modern contraceptives may be greater for those living with HIV, particularly if access to HIV services in interrupted during the migration process. Future research should consider exploring differentiated integrated family planning and HIV services for mobile populations in sub-Saharan Africa, including dual HIV prevention and family planning methods.

## Data Availability

All data produced in the present study are available upon reasonable request to the authors

## Acknowledgements

We thank the Rakai Health Sciences Program as well as the participants of the Rakai Community Cohort Study who made this study possible. We appreciate the help of Dr. Tom Lutalo We also thank Joseph Ssekasanvu and Xinyi Feng who supported the biostatistical analyses presented in this manuscript. This study was supported by the National Institute of Allergy and Infectious Diseases (R01AI087409, U01AI075115, U01AI100031), the National Institute of Child Health and Development (R01HD050180, R01HD070769), the National Institute of Mental Health (F31MH095649, R01MH099733), and the National Institutes of Health Fogarty International Center (D43TW010557). The funders had no role in study design, data collection and analysis, decision to publish, or preparation of the paper.

## Abbreviations

HIV: Human immunodeficiency virus
ART: Antiretroviral therapy
RCCS: Rakai Community Cohort Study

---

**Supplemental Table 1.** Factors associated with unsatisfied contraceptive demand among 3,417 sexually active female participants in the RCCS with no intention of getting pregnant in the next year – sensitivity analysis excluding condoms.

|  | n/N (%) | Unadjusted PR<br>[95% CI] | p-value | Adjusted PR<br>[95% CI] | p-value |
| --- | --- | --- | --- | --- | --- |
| **Migration status** |  |  |  |  |  |
| Resident | 1595/2822 (56.52%) | Ref |  | Ref |  |
| In-migrant | 358/595 (60.17%) | 1.06 [0.99, 1.15] | 0.093 | 1.06 [0.98, 1.16] | 0.163 |
| **HIV serostatus** |  |  |  |  |  |
| HIV-seronegative | 1609/2826 (56.94%) | Ref |  | Ref |  |
| HIV-seropositive | 344/591 (58.21%) | 1.02 [0.95, 1.10] | 0.566 | 0.97 [0.89, 1.06] | 0.48 |
| **Interaction term between migration and HIV-positive serostatus** | 75/115 (65.22%) | 1.15 [1.00, 1.31] | 0.050 | 1.12 [0.94, 1.33] | 0.21 |
| **Age** |  |  |  |  |  |
| 15-24 | 588/910 (64.62%) | Ref |  | Ref |  |
| 25-34 | 700/1462 (47.88%) | 0.74 [0.69, 0.80] | <0.001 | 0.83 [0.77, 0.90] | <0.001 |
| 35-49 | 665/1045 (63.64%) | 0.98 [0.92, 1.05] | 0.652 | 1.10 [1.01, 1.20] | 0.018 |
| **Education** |  |  |  |  |  |
| Some primary | 1141/1991 (57.31%) | Ref |  | Ref |  |
| Post primary | 812/1426 (56.94%) | 0.99 [0.94, 1.05] | 0.832 | 1.00 [0.94, 1.06] | 0.931 |
| **Marital status** |  |  |  |  |  |
| Never married | 360/463 (77.75%) | Ref |  | Ref |  |
| Currently married | 1245/2411 (51.64%) | 0.66 [0.62, 0.71] | <0.001 | 0.67 [0.62, 0.72] | <0.001 |
| Previously married | 348/543 (64.09%) | 0.82 [0.76, 0.89] | <0.001 | 0.79 [0.72, 0.87] | <0.001 |
| **SES** |  |  |  |  |  |
| Lowest | 283/444 (63.74%) | Ref |  | Ref |  |
| Low middle | 590/1016 (58.07%) | 0.91 [0.83, 0.99] | 0.037 | 0.92 [0.84, 1.00] | 0.060 |
| High middle | 577/979 (58.94%) | 0.92 [0.85, 1.00] | 0.079 | 0.94 [0.86, 1.02] | 0.155 |
| Highest | 501/976 (51.33%) | 0.81 [0.73, 0.88] | <0.001 | 0.86 [0.78, 0.95] | 0.002 |
| **Religion** |  |  |  |  |  |
| Other | 13/24 (54.17%) | Ref |  | Ref |  |
| Catholic | 1267/2230 (56.82%) | 1.05 [0.72, 1.51] | 0.800 | 1.12 [0.76, 1.67] | 0.565 |
| Protestant/Pentecost | 417/704 (59.23%) | 1.09 [0.75, 1.59] | 0.639 | 1.17 [0.78, 1.74] | 0.443 |
| Muslim | 256/459 (55.77%) | 1.03 [0.71, 1.50] | 0.879 | 1.11 [0.74, 1.66] | 0.602 |
| **Occupation** |  |  |  |  |  |
| Agriculture/housework | 1137/1997 (56.94%) | Ref |  | Ref |  |
| Non agriculture | 816/1420 (57.46%) | 1.00 [0.95, 1.07] | 0.758 | 0.94 [0.89, 1.00] | 0.071 |

PRR=Prevalence risk ratio; adjRR=adjusted prevalence risk ratio; 95% CI=95% confidence interval; Ref=reference

